# Impact of COVID-19 on Public Research Recruitment

**DOI:** 10.1101/2020.07.21.20158956

**Authors:** Yichuan Grace Hsieh, Holly Parker, Greg Estey, Stephen Lorenz, Mark Wylie, Xiaofeng Zhang, Joseph Lopiccolo, Lloyd Clarke, Jeanhee Chung

## Abstract

The coronavirus disease 2019 (COVID-19) pandemic created a major challenge for clinical trials recruitment as early attention was focused on matters of public health and clinical care, and research--outside of COVID-19-- essentially shut down. Rally with Partners (rally.partners.org), an Internet-based portal for clinical research volunteer recruitment, continued to support studies that continued their recruitment during this period and additionally, implemented several measures to support COVID-19 research. In this paper, we summarize our experiences and preliminary results.

## Introduction

The coronavirus disease 2019 (COVID-19) pandemic created a major challenge for clinical trials recruitment as early attention was focused on matters of public health and clinical care, and research--outside of COVID-19-- essentially shut down. Rally with Partners (rally.partners.org), an Internet-based portal for clinical research volunteer recruitment, continued to support studies that continued their recruitment during this period and additionally, implemented several measures to support COVID-19 research. Rally continued to track visitors and volunteer sign-up across all recruiting studies and offers one lens through which to view public research recruitment during this period. This paper summarizes our experiences and preliminary results.

## Methods

The Rally web site is implemented using the programming language Java and application frameworks that support the creation of responsive (mobile-friendly) web applications, with a Microsoft SQLServer database as the repository for research study postings. Recruitment services and progress tracking based on the site are provided to both study teams and potential research recruits. The software development methodology and support processes are intended to put the software development team close to end-user demands, enabling rapid comprehension and accommodation of changes in end-user priorities.

The following changes to the Rally site and support processes were made to support COVID-19 research:

1. Released a focused COVID-19 page with specially curated COVID-19 studies on April 1, 2020. Studies recruiting from the public (“open to public”) and in the hospital setting (“by invitation only”) were included
2. Added new workflow for by-invitation-only COVID-19 studies and worked with IRB towards complete coverage of therapeutic studies
3. Integrated with COVID Pass, MGB’s digital symptom screening and attestation tool, to display healthcare worker-targeted research studies on April 23, 2020.
4. Coordinated with Marketing and Communications to include MGB studies in broadcast emails and social media

Rally visitors, studies published in Rally, and potential participant contact submissions (through an “I Am Interested” form) between March 1, 2020 and June 11, 2020 were included in the data analysis. Descriptive statistics was used to summarize the data using the IBM SPSS Statistical Software Version 24.

## Results

### Visitors to Rally and volunteer interest in research

During the study period, more than 84,000 visitors (60,419 in year 2019) visited Rally with weekly visitors ranging from 1,905 to 11,868. Figure 1 showed how these variations correspond to some of the current events. The unique visitor count reached a nadir of 1,905 in mid-late March (March 22-28th). Two hundred and twenty-six non-COVID studies retained a “recruiting” status during this period. The first COVID-19 study was published on Rally on March 21, 2020; there were a total of 28 COVID-19-related studies subsequently published (as of June 9, 2020). Among them, 18 (62%) were open to the public. Among all contacts submitted between March 21 and June 6 (n=9,982), almost three-quarters were submitted to the 17 COVID-19 publicly recruiting studies (Figure 2). More than 25% of contacts were shared with non-COVID-19 studies.

**Figure 1.**
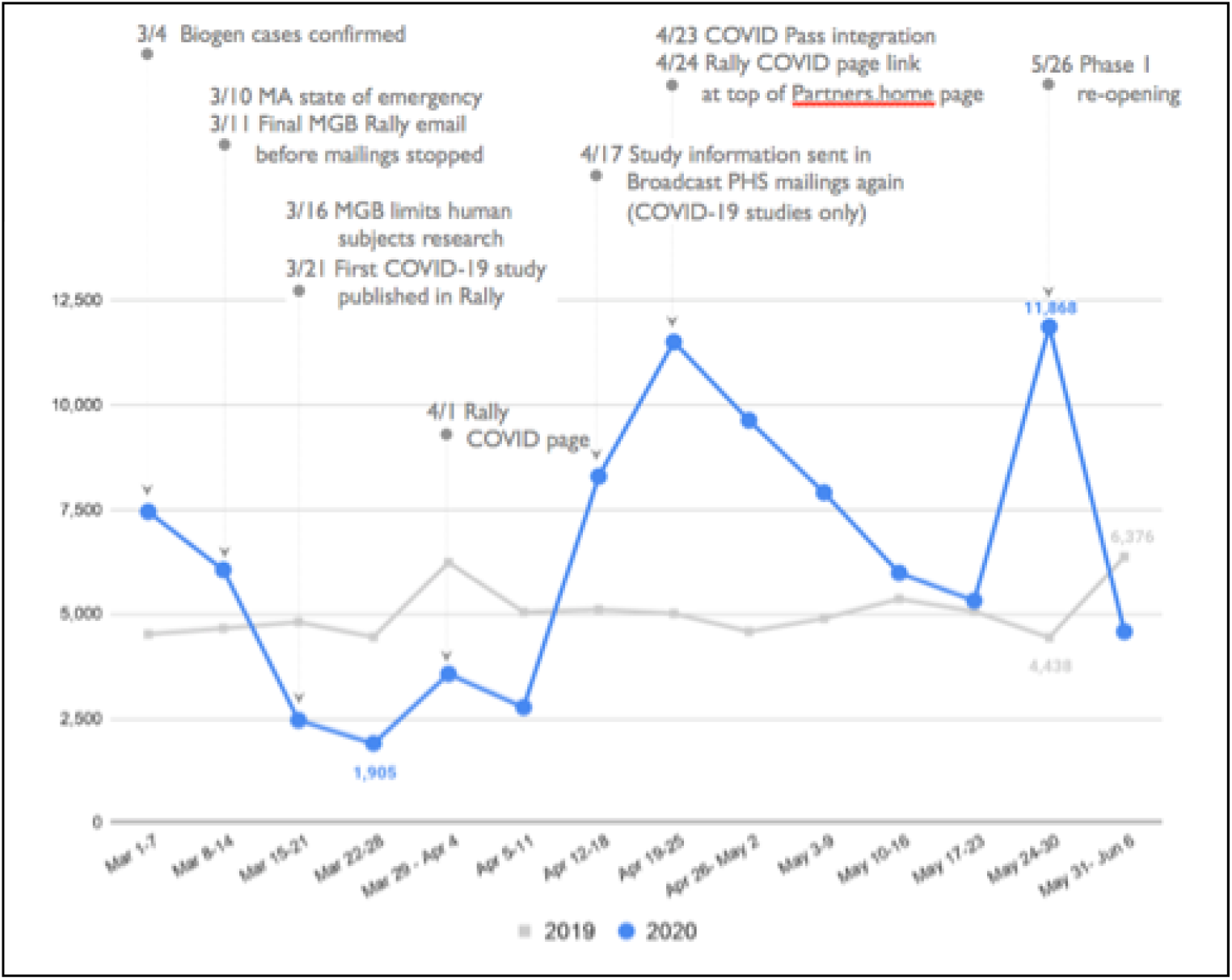
Rally weekly unique visitors.

**Figure 2.**
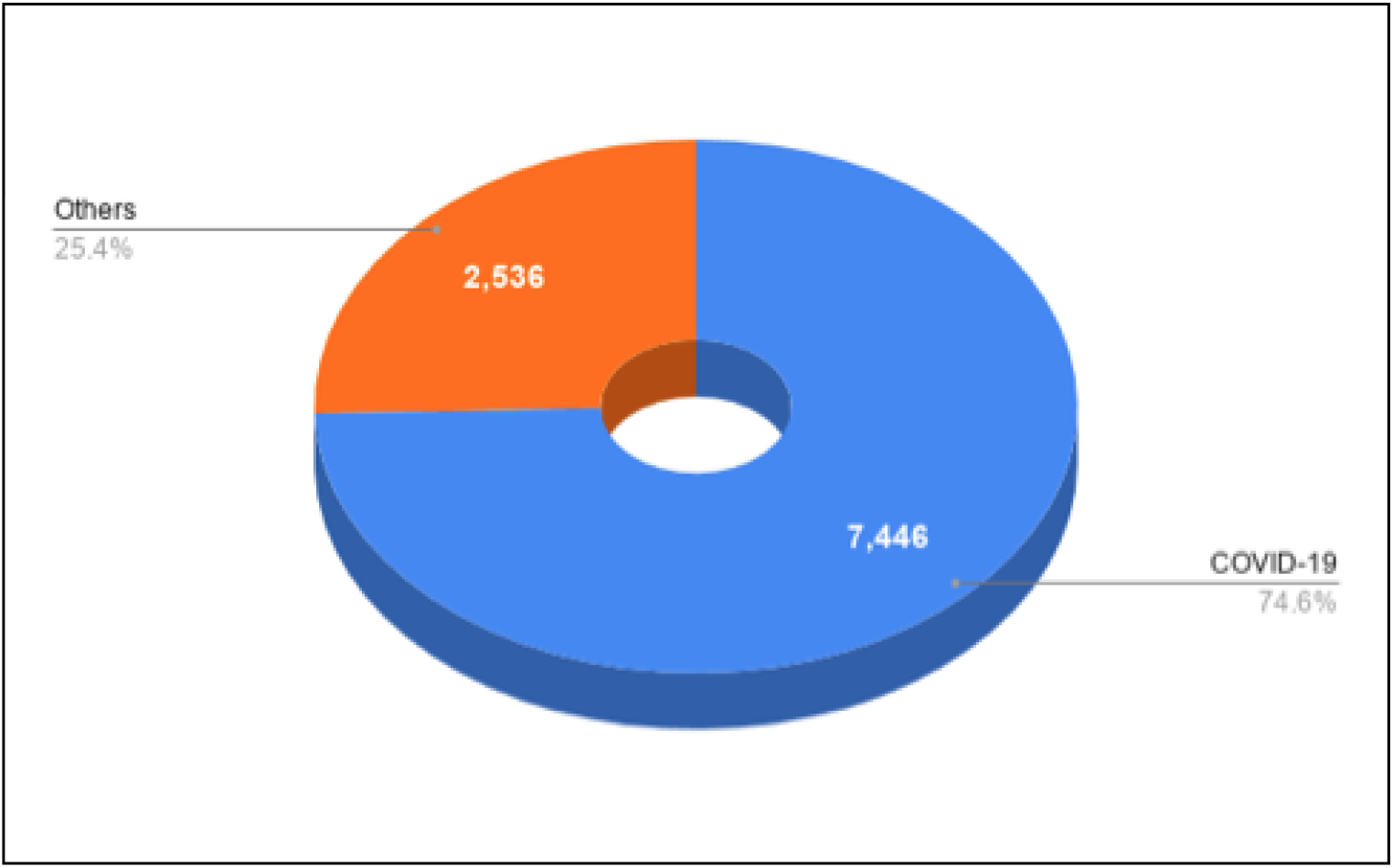
Contact submissions to COVID-19 related studies vs. others.

### Impact of COVID-19-specific campaigns

Rally’s COVID-19 page was released on April 1, 2020. Between April 1 to May 31, this page was viewed more than 9,700 times by 5,700 unique visitors.

Rally integrated with COVID Pass on April 23, 2020. During the first 7-week period after release, almost 9,000 MGB employees from COVID Pass visited Rally with more than 300 contact submissions to 23 studies (more than half of them were COVID-19 related studies).

Another approach was to coordinate with Marketing and Communications to include MGB studies in various broadcast emailing and social media. The first broadcast emails were sent out on April 17, 2020 via Broadcast PHS. Figure 3 showed various recruitment channels of contact submissions during the first 8-week. About 48% (n=3,887) of them came from a deliberate recruitment channel (Broadcast PHS, Weekly Public, Broadcast MGH, Mass General Research Institute, or BWH Research Institute).

**Figure 3.**
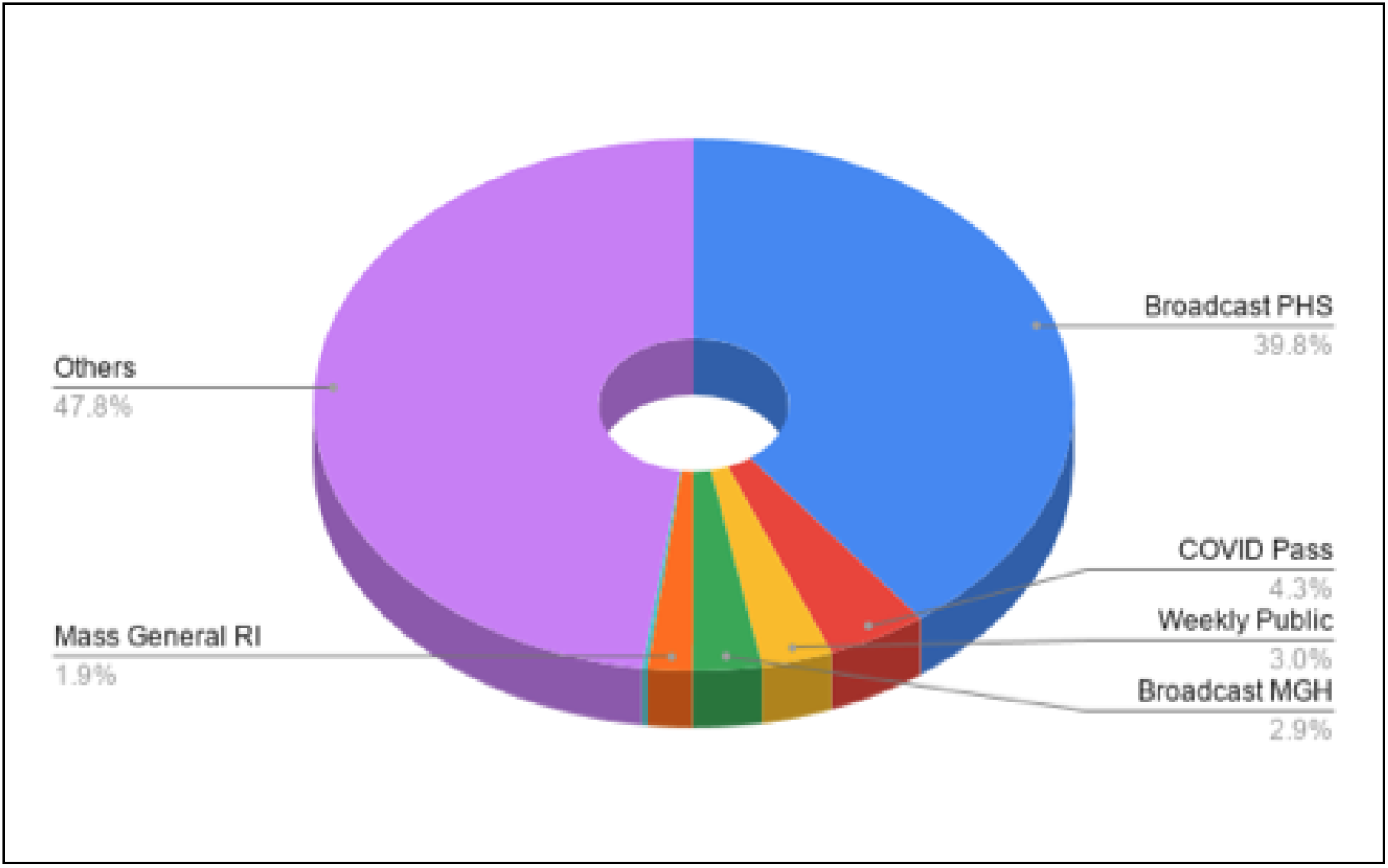
Recruitment channel of contact submissions (n=8,116); duration: 4/17/2020-6/11/2020.

## Conclusion

Volunteer interest in research plummeted during the early phases of the pandemic, but rallied again once COVID-19 research studies began recruiting. Overall, the number of visitors to Rally was higher compared to the same period in 2019. Although COVID-19 research dominated public interest during our study period, some interest continued for non-COVID-19-related research. Recruitment channels played a vital role to reach potential study volunteers. Interest via emails (more than half of overall contact submissions) reflects not only employee engagement, but also the engagement of their extended community with whom these messages have been shared (e.g., one Facebook posting on May 24, 2020 has generated more than 5,000 visitors to Rally in 3-day period). We cannot be certain, at this time, if the increase in research interest arose from new populations as a consequence of the pandemic, or if this research interest will last, but it represents an opportunity to more vigorously engage a public that may now better understand the role of research in their lives.

## Data Availability

aggregated data will be available upon request

